# Controls-Only Quality Control Metric from Early GWAS Can Attenuate Gene-Sex Interaction Signals in Contemporary Large-Scale Studies

**DOI:** 10.64898/2026.07.22.26358697

**Authors:** Desmond Zeya Chen, Marla Mendes de Aquino, Clement Ma

## Abstract

The gene-sex association quality control (QC) metric was developed during the era of early small-sized genome-wide association studies and typically filter variants based on controls alone. While this practice had minimal impact in early studies, in contemporary large-scale settings they can introduce systematic bias by disproportionately discarding variants with pronounced sex differences in allele frequencies (AF), potentially removing real gene-sex interaction signals.

To address this limitation, we introduce Sex-Prevalence Adjusted Allelic Difference Estimates (SPADE) metric, an X chromosome-inclusive QC framework that not only preserves potentially informative variants exhibiting gene-sex interaction but also enables a rapid and exploratory scan for such interaction. SPADE adjusts for sex-specific disease prevalence, maintains correct type I error control, and reduces the risk of falsely excluding variants compared to existing QC approaches. Extensive simulations across diverse disease architectures demonstrated that SPADE remained well calibrated, whereas the controls-only approach exhibited massively inflated type I error when variants were associated with disease. We further developed an open-source command-line software implementation to facilitate its application in large-scale genetic studies.

Applying SPADE to an autism spectrum disorder (ASD) case-control cohort (6,873 cases and 8,981 controls) comprising of the Autism Speaks MSSNG, Simons Simplex Collection (SSC), and Simons Powering Autism Research (SPARK) datasets, we demonstrate that SPADE is well calibrated relative to a controls-only approach. Notably, the complementary exploratory gene-sex interaction scan implicates a sex antagonistic region encompassing *RBMX2, SLC25A14*, and *BCORL1* (lead SNP rs150885581: A>G, p = 6.19 × 10^-9^), providing candidate genes for future functional investigation.

## Introduction

With the rapid advancement of sequencing technologies and the accompanying reduction in cost, ever-increasing sample sizes in genome-wide association studies (GWAS) have enabled the discovery of genetic loci at finer resolution and provided deeper insights into disease pathogenesis^1^. However, this increase in scale may also amplifies subtle biases in statistical procedure, leading to biased inferences. More specifically, a standard GWAS quality control practice is to filter out genetic variants that exhibit pronounced differences in allele frequencies between males and females^2-6^. This step, based on the sex differences in minor allele frequency (sdMAF) metric^7^, is designed to remove technical artifacts and prevent spurious associations in downstream analyses. However, this practice rests on a fundamental and often unexamined assumption: that systematic sex differences in genotype data are more likely to be technical noise rather than biology. When sex is a fundamental modifier of disease risk, it remains unclear to what extent the pursuit of data cleanliness comes at the expense of discarding variants that encode sex-specific disease mechanisms.

This concern is especially relevant given the pervasive role of biological sex in shaping disease susceptibility and clinical outcomes. Biological sex differences arise from the interaction of genetic, hormonal, developmental, and environmental factors^8-11^. These differences contribute to distinct physiological, cognitive, and behavioral traits between males and females, affecting health outcomes, disease susceptibility, physical performance, and psychological tendencies^12-15^. These differences are especially pronounced in neuropsychiatric disorders. Females are approximately twice as likely to develop depression and anxiety disorders, whereas males are diagnosed more frequently with attention-deficit/hyperactivity disorder and autism spectrum disorder (ASD)^16,17^. In particular, ASD exhibits a marked male predominance (≈3.8:1), providing strong evidence for sex-dependent disease mechanisms and motivating the search for gene-sex interactions^18^. This has motivated increasing interest in identifying gene-sex interactions genome-wide. Despite this, the X chromosome, which harbors numerous genes involved in neurodevelopment, remains analytically challenging because of differences in ploidy, X-chromosome inactivation, and non-pseudoautosomal region (NPR) inheritance, complicating statistical modeling^19,20^.

However, limitations in detecting gene-sex interactions are not confined to statistical testing methods but also arise from upstream quality control procedures that determine which variants are retained for downstream analysis. In case-control GWAS, general population gene-sex independence assumption is operationalized by testing gene-sex association exclusively in controls^2,3,21,22^. As we show later in the Methods, such artifacts can also induce spurious signals in gene-sex interaction analyses for sex-biased diseases if left unaddressed. Moreover, the procedure might be suboptimal, as it relies heavily on the rare disease assumption, under which controls are treated as a representative proxy for the general population. In practice, this assumption often fails. Both Gatto *et al*. and Pierce *et al*.^23,24^ demonstrated that control samples may not accurately reflect population-level distributions, even when disease prevalence is low. Similarly, controls-only Hardy-Weinberg equilibrium (HWE) filtering may inadvertently remove true susceptibility SNPs, as genuine risk variants are expected to deviate from HWE in cases^25^. Accordingly, Khramtsova *et al*. recommended flagging markers for further scrutiny, rather than removing them outright^2^.

As GWAS sample sizes grow, loci exhibiting sex differences in controls are more likely to be observed and mistakenly removed, though some may reflect genuine gene-sex interactions. For example, consider a disease with 1% prevalence, no sex differences in the underlying population, and no other confounding effect, such that the population-level gene-sex odds ratio is (9900/100)/(9900/100) = 1. Despite this, sex differences can readily emerge among controls ((9815/85)/(9865/35) ≈ 0.41, p < 10^−5^), purely due to conditioning on disease status. Meanwhile, the ratio of sex-specific odds ratio, capturing gene sex interaction, is 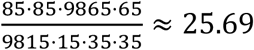, with *p < 10*^*−15*^, indicating a strong interaction effect. This example highlights a fundamental tension between data cleaning and discovery: the tools we use to remove technical noise can inadvertently erase true biological signals.

To overcome these limitations, we present the Sex-Prevalence Adjusted Allelic Difference Estimates (SPADE) metric, an X chromosome-inclusive QC framework that not only preserves potentially informative variants exhibiting gene-sex interaction but also enables a rapid and exploratory scan for such interactions. Our core insights are: (1) with sex penetrance, we can generate general population AF estimates and therefore perform a more robust inference, (2) AF differences between sexes conditioning on affection status, which previously may be treated as technical noise in quality control pipelines, actually provide direct evidence of biological gene-sex interactions when properly contextualized.

In this paper, we: (1) formalize the statistical foundation of SPADE and demonstrate its validity through simulations; (2) develop an extended QC statistic for combined case-control data to substitute the existing controls-only QC; (3) extend the case-only framework by combining evidence from cases and controls using Fisher’s method; (4) demonstrate the utility of SPADE by applying it to ASD GWAS data; and (5) provide a user-oriented and efficient software under command-line interface. By bridging quality control and biological inference, SPADE offers a principled, robust, and practical solution for uncovering the genetic basis of sex differences in phenotype.

## Methods

### Bias of Allelic Differences by Sex in Controls

Let *Y*_*i*_ ∈ {*a, u*} denote the disease status of individual *i* among *n* samples, with *Y*_*i*_ = *a* ≡ 1 indicating a case and *Y*_*i*_ = *u* ≡ 0 indicating a control. Let *G*_*i*_ denote the genotype at a bi-allelic SNP, typically coded additively as the number of minor alleles, *G*_*i*_ ∈ {0,1,2} . This coding applies to all females genome-wide and to males for autosomal and pseudoautosomal region (PAR) variants. For variants in the X chromosome non-pseudoautosomal region (NPR), hemizygous males are coded as *G*_*i*_ ∈ {0,1} when no dosage assumption is made, or as *G*_*i*_ ∈ {0,2} under the assumption of random, complete X chromosome inactivation (XCI) in females^20^. Throughout, we adopt the {0,1} coding without loss of generality. Let *S*_*i*_ ∈ {*f, m*} denote biological sex (*f* = female ≡ 0, *m* = male ≡ 1), where *π*_*s*_ and *p*_*s*_ are correspondingly the sex-specific disease prevalence and allele frequencies (AF). *p*_*s*_ can be either the major or minor allele frequency, as we are only interested in sex differences, and the selection will not affect the magnitude of the difference. *p*_*s*,*y*_ is the sex-specific AF conditioned on disease status. Sex-specific penetrance is defined as *κ*_*g*,*s*_ . Under a multiplicative model, *κ*_*g*,*s*_ = *k*_0,*s*_ *⋅* η_*g*,*s*_, where *k*_0,*s*_ is the baseline penetrance (for *g* = 0) and η_*g*,*s*_ is the genotype specific multiplicative factor, allowed to differ by sex. Finally, let *δ*_*s*_ denote the sex-specific deviation from HWE. The autosomal/PAR major AF in cases for sex *s* is (full derivation see Note S1):

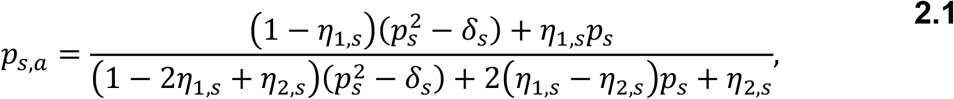

and the special case for NPR males follows:

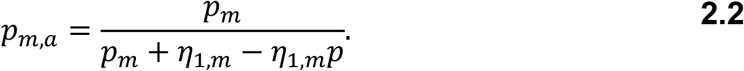

Then the allelic difference by sex in controls is:

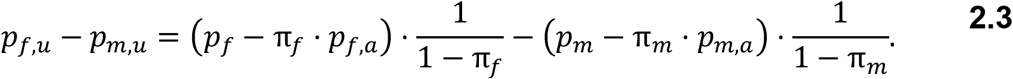

The existing gene-sex association in controls metric assumes sex-specific prevalence is rare and *π*_*s*_ can be replaced with 0, where equation 2.3 reduces to *p*_*f*_ *− p*_*m*_, the general population allelic differences. Under the null hypothesis (*H*_0_: *p*_*f*_ = *p*_*m*_), *p*_*f*,*u*_ *− p*_*m*,*u*_ should ideally equate to 0, we therefore use the term bias for it. The bias can interfere with the quality control procedure in larger sample sizes. The existing controls-only metric have conditioned on disease status; hence, it is effectively a “case-only design”, of which, could pick up genuine gene-sex interaction signals instead^26^.

### Sex-Prevalence Adjusted Allelic Difference Estimates Metric

We propose an alternative approach by testing the sex prevalence adjusted allelic difference which estimates the general population difference compared to controls-only quantity. We let 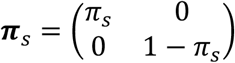 a diagonal matrix of disease prevalence and its complement. Given the availability of reliable external prevalence estimates, we consider *π*_*s*_ fixed throughout. Define **1**_**2**_ = (1, 1)^⊤^ and *p*_*s*_ = (*p*_*s*,*a*_, *p*_*s*,*u*_)^⊤^, a vector of AF in cases and controls for sex *s*.

Hence, by Bayes’ rule the population-level sex consistency hypothesis *H*_0_: *p*_*f*_ = *p*_*m*_ can be expanded as *H*_0_: **1**_**2**_^⊤^*π*_*s*_*p*_*f*_ = **1**_**2**_^⊤^*π*_*s*_*p*_*m*_.

We further define a sex specific covariance matrix of estimated AF, 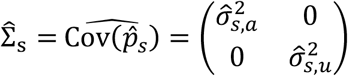, then the proposed QC metric is:

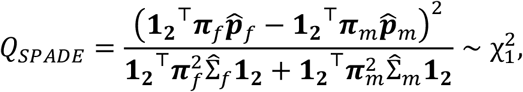

Here, 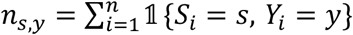 is the sex by affection status strata-specific sample size and 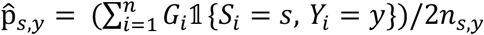 in general, and 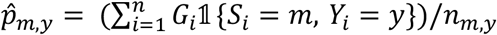 specifically for male NPR. The variance 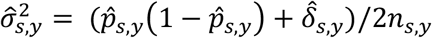 for autosomal/PAR variants and 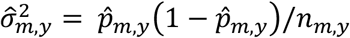 for male NPR.

In large sample settings, each sample AF converges to the true AF. By Slutsky’s theorem the unsquared numerator will converge to *p*_*f*_ − *p*_*m*_ with mean 0 under the null hypothesis. Then, the Central Limit Theorem yields an asymptotic Chi-square distribution with 1 degree of freedom for the test statistic. The variance, however, is based on normal approximation, where it is valid under sufficiently large sample sizes, typically when all expected cell counts are *≥* 5 across four strata. When this condition is violated, the asymptotic approximation may be unreliable.

### Rapid and Exploratory Scan of Gene-Sex interaction via Fisher’s Combination Function

In conjunction with the QC test, we also propose a quick gene-sex interaction scan as a complement. Since both case-only allelic test by sex and controls-only allelic test by sex can potentially detect gene-sex interaction^13^, and with the assumption that cases and controls are comprised of separate cohort with independent sampling procedure, we can pool the corresponding case-only p-value (*P*_*case*_) and controls-only p-value (*P*_*ctrl*_) via Fisher’s combination function^27^ as the following: 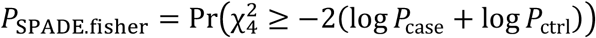.

### Simulation Methods

We assessed the Type I error (T1E) rate of SPADE against controls-only allelic test by sex across 3 sample size settings reflecting: early GWAS (n=8,000), contemporary GWAS (n=40,000), and large-scale cohort (n=400,000) (Additional details in Note S2). Genotype/haplotype counts of males, females for both cases and controls were multinomially random sampled with respect to the following data generating mechanism:

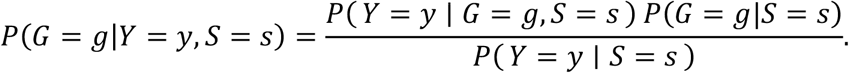

The denominator is either Π_*s*_ or (1 − Π_*s*_), while *P*(*G* = *g*|*S* = *s*) can be derived from MAF (*p*) and HWE delta (*δ*). For autosomes and the PAR region (with *G* ∈ {0,1,2}), the genotype probabilities are given by (*p*^2^ *+ δ*, 2*p*(1 − *p*) − 2*δ*, (1 − *p*)^2^ *+ δ*). For male NPR variants on the X chromosome (with *G* ∈ {0,1}), the corresponding probabilities are (1 − *p, p*).Sex-specific penetrance *P*(*Y* = 1 ∣ *G* = *g, S* = *s*) is modeled multiplicatively, where each of the conditional penetrance *P*(*Y* = 1 ∣ *G* = *g, S* = *s*) = *κ*_0,*s*_ *⋅* η_*g*,*s*_, can be interpreted as a multiple of the baseline penetrance and the baseline penetrance is 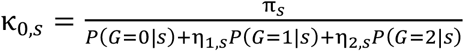 . For males’ NPR, we set η_2,*s*_ = 0 . With *p, δ*, η_1,*s*_, η_2,*s*_ and sex-specific sample size pre-specified, we can simulate genotype counts for both male and female cases. Moreover, sex-specific non-penetrance *P*(*Y* = 0 ∣ *G* = *g, S* = *s*) = 1 − *P*(*Y* = 1 ∣ *G* = *g, S* = *s*) is used to simulate controls.

For effect modeling, we considered four genetic architectures: (strong main effect with minor interaction, male-only effect, female-only effect, and strong interaction with minor main effect) for NPR variants and three for autosomal variants (Table S1). Three AF were chosen (0.01, 0.1, 0.35) to reflect rare, common, and high frequency variants. Each scenario was replicated 100,000 times across a range of female-male prevalences ((1%,3%), (4%,6%), (9%,11%), (19%,21%), (29%,31%)). A significance threshold of 1×10^-4^ was used for type I error (T1E) rate comparison. Furthermore, four representative scenarios were considered to illustrate the distribution of p-values, defined by sample size (40,000 or 400,000) and sex-specific disease prevalence (3% in males and 1% in females, or 16% in males and 14% in females). The AF was fixed at 0.1, and simulations were conducted under a consistent strong main-effect genetic architecture. For each scenario, 10,000 simulation replicates were performed. Power simulation set up can be found in Note S2.

### Application to ASD cohort

We applied the SPADE to X chromosome NPR summary output from Mendes et al.^17^, whose analysis motivated the present work. ASD case data comprised 6,873 individuals (5,639 males and 1,234 females) from three whole-genome sequencing cohorts: MSSNG^28,29^, Simons Simplex Collection (SSC)^30^, and SPARK^31^. Control data included 8,981 individuals (3,911 males and 5,070 females) from the Medical Genome Reference Bank (MGRB)^32^ and the CanCOGeN HostSeq initiative^33^. Variant and Sample-level QC were performed by the original investigators. All ethical approvals and participant consents were managed by the data providers. Controls-only sdMAF summary statistics was used as a comparison; additionally, a Fisher’s exact test^34^ of allele by sex was also performed to the controls as a sensitivity analysis. Sex-specific autism prevalence estimates (1.14% in females and 4.3% in males) were obtained from Maenner et al.^18^

Additionally, SPADE.fisher test was applied by combining case and control specific sdMAF summary output. We then compared it against the sex-stratified heterogeneity test results generated from the GWAMA^35^ tool (in output column “gender_heterogeneity_p.value”). Such test is done by computing 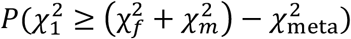 here, 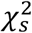 are sex-stratified chi-square summary statistics of betas from logistic regression models adjusted for covariates, and 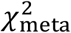 is the inverse variance weighted fixed effect meta-analysis of sexes. LocusZoom plots were generated using My LocusZoom web application^36^ and LDlink^37^(v7.0.0), with linkage disequilibrium estimated from the 1000 Genomes Project^38^ European samples (GRCh38 High Coverage).

### Software Implementation

SPADE was implemented as an R-based command-line tool. It accepts PLINK-formatted genotype count data and supports analyses of both pseudo-autosomal (PAR) and non-PAR variants by automatically applying the appropriate statistical procedures according to genomic context. The software performs input validation, variant filtering, statistical estimation, and logging to ensure reproducible analyses. A command-line interface was developed using the ‘argparse’ R package^39^, and genome-wide analyses can be parallelized at the chromosome level.

## Results

### Calibration and statistical properties of SPADE

We assessed T1E rate of SPADE versus controls-only allelic test by sex across a range of scenarios. As expected, in early GWAS setting with n=8,000, both SPADE and controls-only variants by sex association maintained good calibration across genetic structures, allele frequencies, genome region and sex prevalence ratios (Figures S1-S6). In this setting, controls-only QC statistics were constructed using 4,000 controls, resulting in limited power to detect the bias. When less stringent significance thresholds (e.g., *α* = 0.01 or *α* = 0.05) were applied, statistical power increased, which resulted in inflated T1E for controls-only QC. In contrast, SPADE remained good calibration across all settings.

However, for contemporary GWAS and large biobank settings, bias caused by sex-disparities in disease emerges and inflates T1E of controls-only QC (Figure 1). The magnitude of the bias increases monotonically relative to disease prevalence or minor allele frequencies alone. The direction is determined by the sex specific genotypic effect size and whether major or minor AF was used for calculating the difference. The inflation in controls-only QC typically increases with respect to the magnitude of the bias, and the impact in the biobank setting is generally higher than that in contemporary GWAS sample size (Figure 1A and S7A). A ten-fold inflation of type I error was observed for common variants (AF = 0.1) at sex-combined prevalence as low as 5% across both sample size settings. In contrast, for rare variants (AF = 0.01), higher prevalence was required to achieve a comparable level of inflation. As prevalence increased, type I error rates approached 1 at *α* = 1 × 10^-4^, indicating that nearly all replicates were rejected under these conditions. In contrast, results from SPADE are well-controlled and invariant to sample size.

**Figure 1:**
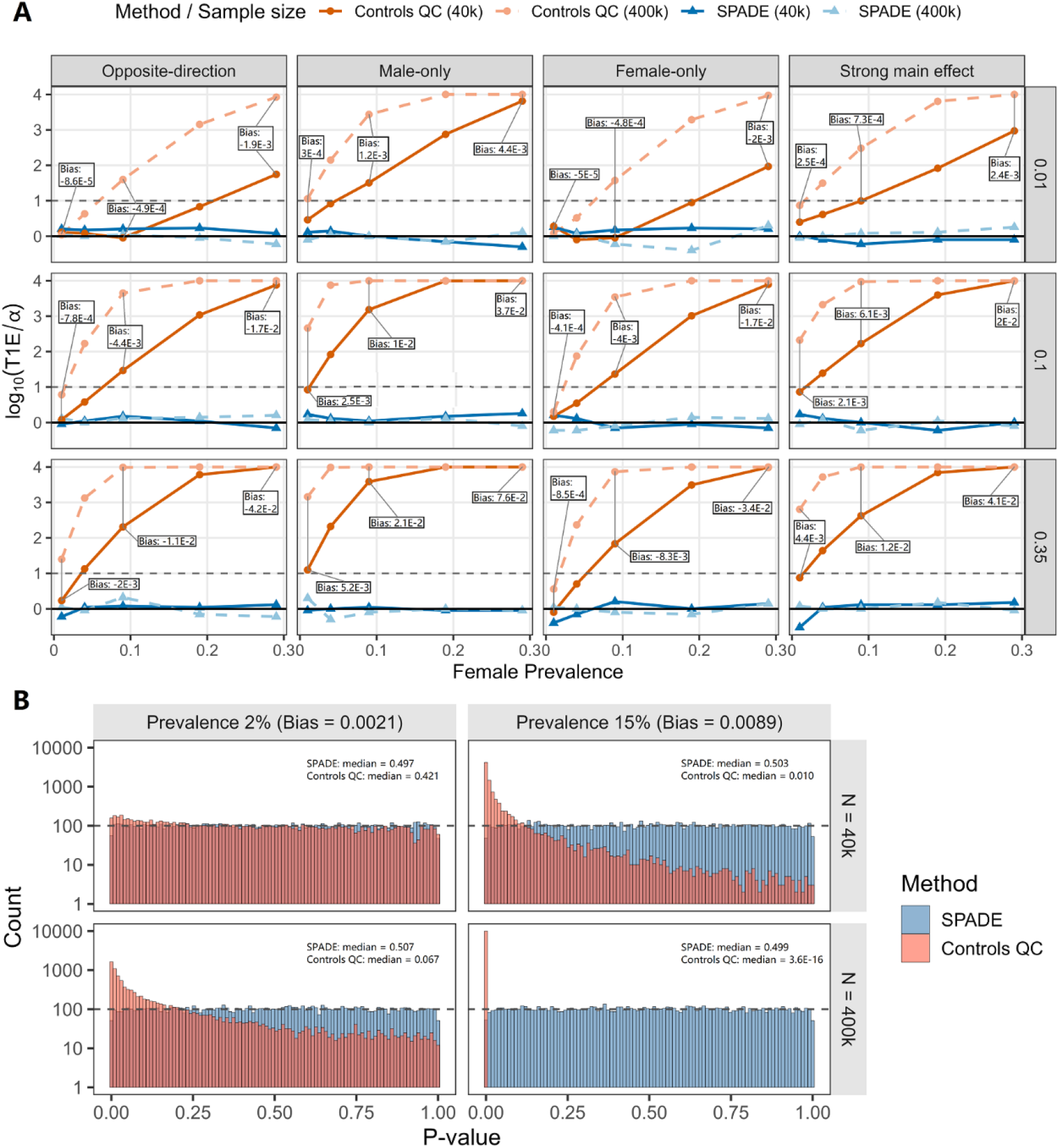
Simulation-based type I error rate (T1E) for SPADE and controls-only QC for chromosome X non-pseudoautosomal region variants. ***(A)*** T1E relative to the nominal *α* = 1 × 10^−4^ across different genetic architectures (columns) and allele frequencies (rows). The gray dashed line at *y* = 1 represents a 10-fold inflation (i.e. T1E = 1 × 10^−3^), while the zero line represents no inflation. **(B)** Empirical p-value distribution of SPADE and controls-only QC across different disease prevalences (columns) and sample sizes (rows). Variants were simulated with AF = 0.1 and under a strong main effect genetic architecture. The gray dashed line represents expected density for a uniform(0,1) random variable.

Under rare sex-combined disease prevalence (2%) and smaller sample size (N=40,000), controls-only QC already deviates from the nominal distribution as a small excess of small p-values; the deviation is more pronounced for larger sample sizes (N = 400,000) (Figure 1B). For common disease prevalence (15%) across both sample sizes, controls-only QC exhibited poor type I error control. SPADE remained well-calibrated, with p-value distributions closely matching the expected null distribution (Figure 1B and S7B). In power simulations, under the setting when there exist true sex differences in controls, SPADE and controls-only QC yielded comparable performance (Tables S2 and S3). However, when errors affected cases, SPADE maintained high power whereas controls-only QC power approached zero.

### Comparison with existing approaches in ASD cohort

We applied SPADE to the ASD cohort from Mendes et al.^17^, and compared it against controls allelic test by sex results. First, controls QC identified 5 SNPs with p-values < *α* = 5 × 10^−6^ across the X chromosome (Figure 2). These signals are subsequently subject to QC filtering, motivating comparison across methods.

**Figure 2:**
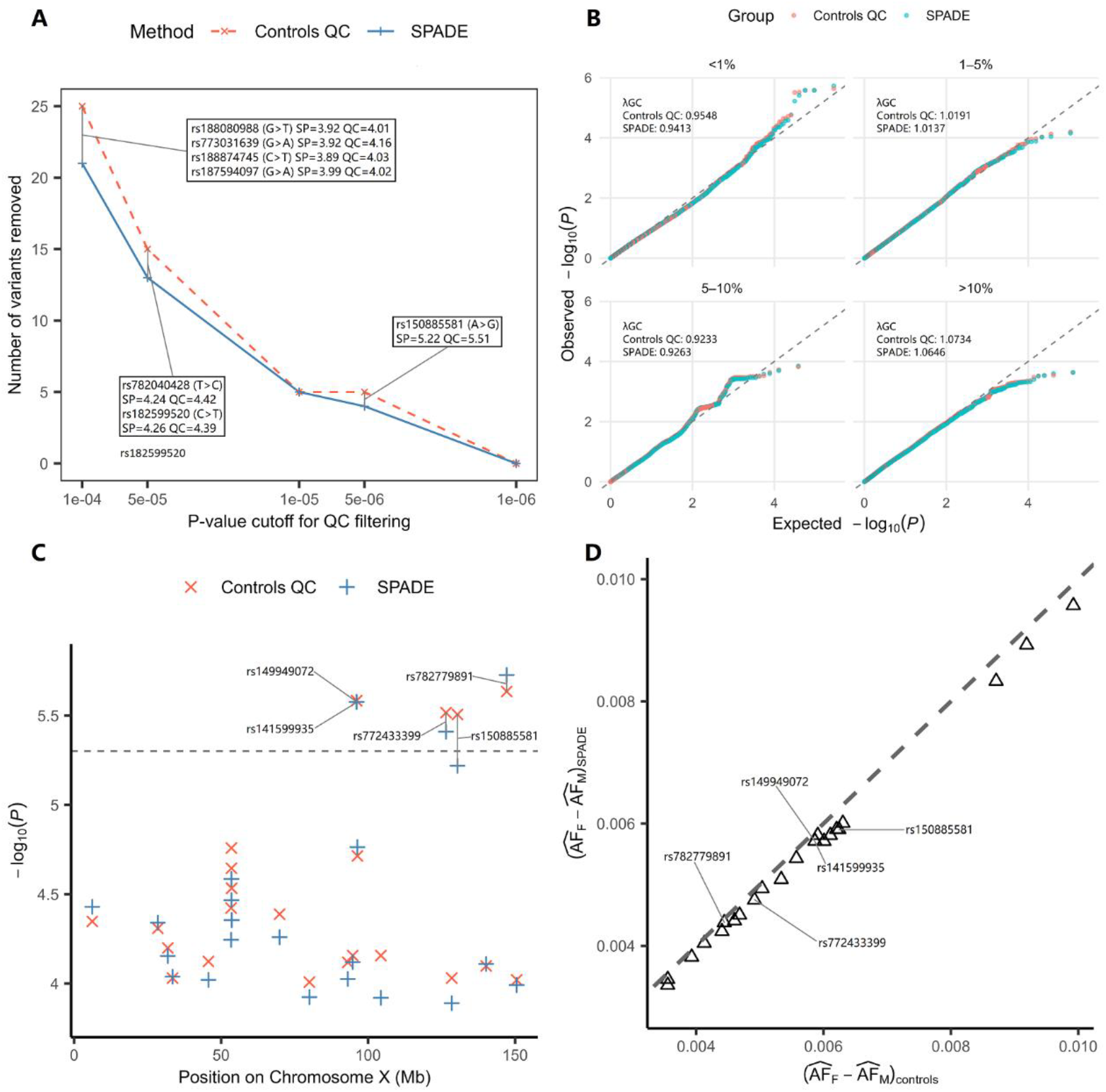
Comparison of SPADE and controls-only QC in the ASD cohort. ***(A)*** Comparison of variant filtering between SPADE and controls-only QC across QC thresholds. Annotated variants indicate SNPs removed by controls-only QC but retained by SPADE. SP and QC denote the −log10(p) from SPADE and controls-only QC, respectively. ***(B)*** Minor allele frequency-stratified quantile-quantile plots by method. The gray dashed line denotes the identity function. Minor allele frequencies were calculated from sex-combined controls. *λ*GC denotes the genomic control inflation factor. ***(C)*** Stacked Manhattan plot of SPADE and controls-only QC results for the X chromosome NPR. Only SNPs with controls QC −log10(p) > 4 are shown. The gray dashed-line represents *α* = 5 × 10^−6^. ***(D)*** Comparison of sex-difference effect size estimates from controls-only QC and SPADE for the SNP subset in subpanel C. The gray dashed-line represents the identity function.

We compared the number of variants removed across a range of QC significance thresholds between SPADE and controls-only QC (Figure 2A). Across all QC significance thresholds, SPADE removes the same or fewer variants compared to controls-only QC. The difference between methods becomes more prominent at higher p-value cutoff (i.e. more stringent thresholds for retaining). Annotated variants highlight loci that are removed by controls-only QC but retained by SPADE for specific cutoff.

To assess the robustness of these observations to the choice of statistical test, we conducted a sensitivity analysis using Fisher’s exact test of sex and allele count in controls. Since Fisher’s exact test is known to be conservative^40^, only two SNPs had p-values below *α* = 5 × 10^−6^, and both had already been removed by the variant-level filter in both SPADE and the controls-only QC because of low cell counts. (Figure S8).

We assessed the calibration of both tests stratified by MAF (Figure 2B). Across MAF strata, both SPADE and controls-only QC are well-calibrated overall, with genomic control inflation factors^41^ (*λ*GC) ranging from 0.94 to 1.06. Both methods show slight type 1 error inflation for the MAF < 1% stratum, with controls-only QC exhibiting greater upper-tail inflation than SPADE. In the marginal QQ plot (Figure S9A), both methods are well-calibrated in the bulk and lower tail (*λ*GC = 0.98 for both), whereas the upper tail of controls-only QC shows mild inflation relative to the identity line. For rare variants (minor allele count < 5), both SPADE and controls-only QC exhibit substantial inflation (Figure S9B). In contrast, Fisher’s exact test is conservative, as expected, in both marginal and MAF-stratified QQ plots for controls-only analyses (Figure S9C & S10).

To avoid overplotting, panel C and D of figure 2 displays only SNPs with controls QC − log_10_(*p*) > 4 . Within this subset, the controls-only analysis generally yielded slightly smaller p-values and modestly larger estimated sex-difference effect sizes than SPADE, consistent with the pattern observed in the simulation study.

### Application to ASD cohort reveals sex-differentiated signals

We next applied the SPADE.fisher test to the ASD cohort and compared its performance with a conventional sex-heterogeneity test. In the genome-wide scan, SPADE.fisher identified a genome-wide significant SNP rs150885581 (− log_10_(*p*) = 8.21), whereas the sex-heterogeneity test was not significant (− log_10_(*p*) = 5.23) (Figure 3A). This suggests SPADE.fisher can identify signals that may be attenuated by existing methods.

**Figure 3:**
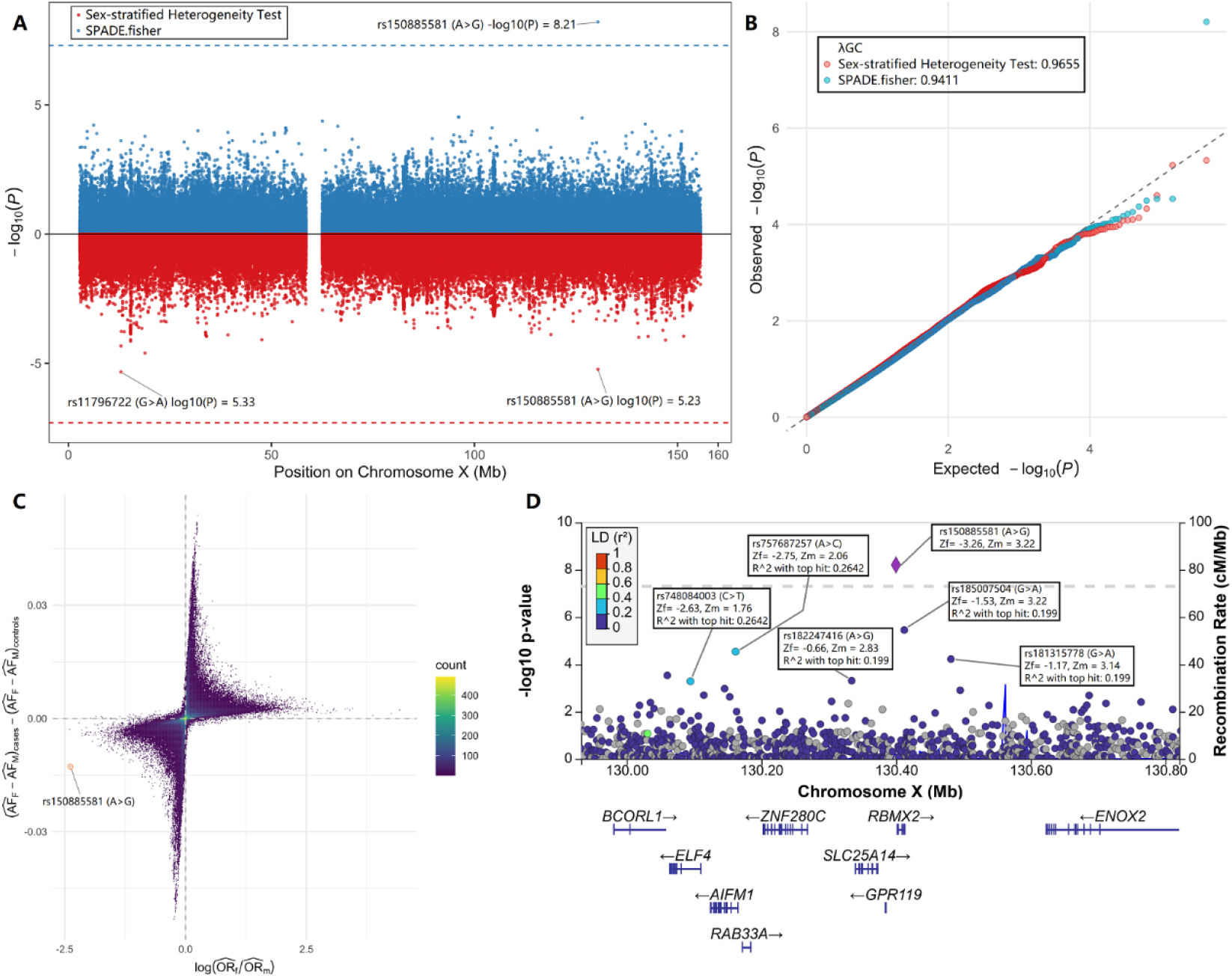
Comparison of gene-by-sex interaction association results in ASD using SPADE-Fisher and sex-heterogeneity tests. ***(A)*** Miami plot of SPADE-Fisher and the sex-heterogeneity tests. Horizontal dashed lines indicate the genome-wide significance thresholds at *α* = 5 × 10^−8^. ***(B)*** Quantile-quantile plots of SPADE-Fisher and the sex-heterogeneity test p-values. The gray dashed line represents the expected null distribution. ***(C)*** Hexbin plot of case-control differences in sex-specific allele frequencies and the log ratio of female-to-male odds ratios. ***(D)*** LocusZoom plot for the top associated variant rs150885581. Text-annotated linkage disequilibrium (LD) is based on European samples from the 1000 Genomes Project, with annotated SNPs manually recolored to reflect LD values obtained from LDlink.

**Figure 4:**
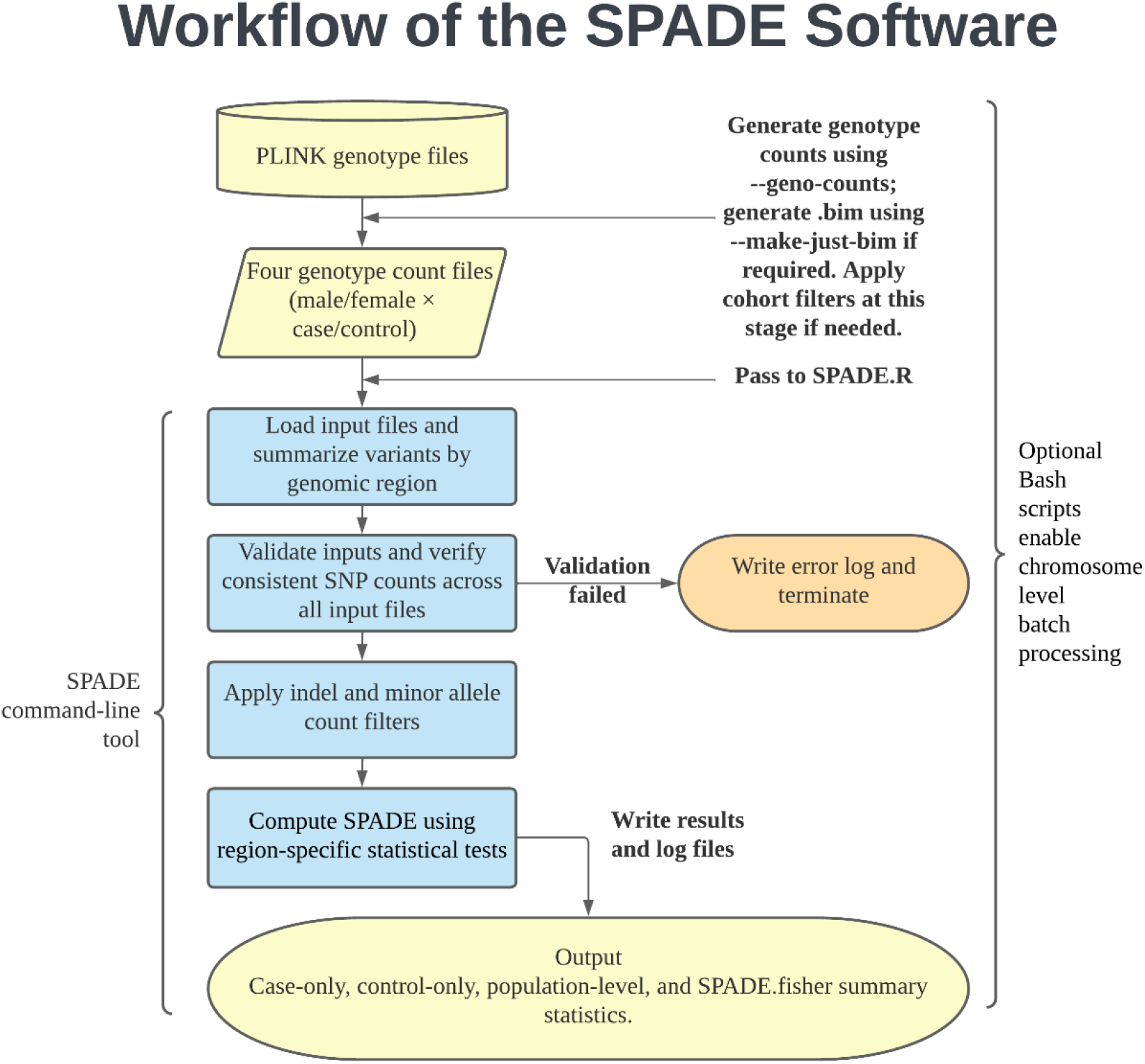
Workflow of the SPADE software. The pipeline illustrates data preparation, input validation, quality control, statistical analyses, and output generation.

Quantile-quantile plots indicated that both approaches showed mild upper-tail deflation with *λ*_*GC*_ < 1.0, consistent with conservative behavior under sparse counts or discreteness of the test statistic (Figure 3B). These findings indicate that the observed genome-wide significant signal was unlikely to be driven by broad residual confounding or uncontrolled test-statistic inflation.

To examine the relationship between alternative formulations of gene-by-sex effects, we compared the case-control difference in sex-specific allele-frequency contrasts against the log ratio of female-to-male odds ratios. Variants were concentrated primarily in the second and fourth quadrants, demonstrating the expected inverse correspondence between these two summaries of interaction (Figure 3C). When variants were further stratified by minor allele frequency, the relationship within strata became approximately linear (Figure S11), indicating that the two statistics capture closely related estimands and differ largely through scaling properties tied to allele frequency.

Locus zoom plot of rs150885581 showed several nearby variants with low linkage disequilibrium showing moderately reduced p-values (though not genome-wide significant) further supports the validity of this association signal. (Figure 3D). Using sex-specific association statistics reported by Mendes et al.^17^, the lead variant displayed opposite directions of effect in females and males, with a negative effect estimate in females and a positive effect estimate in males, consistent with a sex-antagonistic pattern. The associated interval spans *RBMX2, SLC25A14*, and *BCORL1*(Figure 3D).

### SOFTWARE availability

We developed SPADE to provide an engineering realization for allele frequency quality control and the detection of sex-differentiated genetic signals. The software estimates sex-specific allele frequencies and their associated variances and reports multiple statistical tests, including case-only, control-only, and population-level comparisons, together with corresponding effect estimates and significance measures. SPADE supports user-specified case-control samples, and disease prevalence parameters, enabling flexible application across a wide range of study designs. Genome-wide analyses can be performed efficiently using chromosome-level parallelization. Documentation, example workflows, and source code are freely available at GitHub (https://github.com/zeyachen/SPADE).

## Discussion

The expanding scale of GWAS has brought both opportunity and risk: while statistical power increases, so too does sensitivity to subtle biases in quality control. In this study, we demonstrated that a widely adopted QC metric, gene and sex association in control samples, can be systematically biased, leading to either excessive variant exclusion or inflation of inferred effects in contemporary large-scale studies. Through statistical derivation and empirical evaluation, we showed that this bias is not merely a technical nuisance but a reflection of a deeper issue: QC metrics derived from controls alone implicitly assume that the distribution of genetic variants is independent of disease liability, an assumption that becomes increasingly untenable as GWAS sample sizes grow and disease architectures become more complex.

We developed SPADE to address this gap by integrating case and control information to reconstruct population-level allelic distributions, thereby aligning QC with the inferential goals of genetic association testing. The same inferential framework also motivated SPADE.fisher, which extends this perspective to gene-by-sex interaction testing by integrating evidence from case-only, and control-only analyses. A command-line tool is provided to enhance the accessibility and adoption of the SPADE framework in standard genetic association studies. From a methodological standpoint, SPADE introduces a shift in how QC metrics can be conceptualized. Traditionally, QC has been viewed as a diagnostic prelude to inference, a filtering step designed to remove “bad” variants before analyses. Our work suggests that QC metrics themselves can be reframed as inferential tools, capable of revealing biological signal when appropriately parameterized. This diagnostic-to-inferential paradigm extends naturally to other QC dimensions, such as batch effects, population structure, or relatedness, raising the possibility that current best practices may be discarding informative variation under the guise of quality control. In this view, SPADE represents not merely a new algorithm but a template for rethinking how we separate artifact from biology in large-scale genetic data.

Across simulations and application to the ASD cohort, SPADE showed improved calibration while retaining variants that would otherwise be removed under controls-only allelic test by sex QC metric. Observations from real data also revealed SPADE as better calibrated than inflated controls-only allelic test, confirming simulation results. Additionally, the identification of a genome-wide significant locus by SPADE.fisher, but not by the conventional sex-heterogeneity test, suggests that case-only controls-only combined design can potentially be more powerful than conventional case-only design, and heterogeneity test. The near-linear correspondence between allele-frequency contrast statistics and odds-ratio heterogeneity indicates that proposed method and commonly used formulations capture closely related underlying interaction signals, with differences arising primarily from scale and variance parametrization.

The biological findings further reinforce the value of this approach. The observed sex-antagonistic pattern at rs150885581 is notable, as variants with opposite directions of effect between females and males may contribute to sex-differential liability in ASD. The genes *RBMX2, SLC25A14*, and *BCORL1*, all have previously shown to be linked to neurodevelopmental disorders and mental disorders that are comorbid with ASD. The identification of a rare *RBMX2* variant in an ASD proband^42^, together with previous reports linking *RBMX2* to bipolar disorder which shares symptomatic overlap with ASD^43,44^, provides evidence supporting *RBMX2* as a candidate gene for ASD. Reduced expression of mitochondria-related gene *SLC25A14* has been observed in postmortem brain tissue from ASD patients, specifically in the anterior cingulate cortex and motor cortex, suggesting a potential role for this gene in the pathophysiology of ASD^45^. Finally, according to the SFARI Gene database, *BCORL1* is classified as a syndromic ASD gene (Category S)^46^. Collectively, these convergent lines of evidence support the biological plausibility of this genomic region as a candidate locus for further investigation.

Nevertheless, several limitations should be acknowledged. SPADE requires prior knowledge of sex-specific disease prevalence in the general population; inaccurate estimates may lead to biased or misleading inferences. Moreover, SPADE.fisher provides a rapid scan of gene-sex interaction without conditioning on baseline covariates. For secondary confirmatory analyses, *P*_case_ and *P*_ctrl_ can be derived from GWAS of sex in a logistic regression with covariates to fully account for confounding effects. We recognize that the real-data example does not fully capture the ideal conditions for SPADE, where the bias is more pronounced and sample sizes are larger. Future studies applying SPADE to larger cohorts and diseases with higher prevalence will provide a more comprehensive evaluation of its practical performance.

The SPADE framework can also be extended beyond gene-by-sex interaction studies. Because SPADE.fisher builds upon a case-only component, its application to other gene-environment interaction settings requires the standard case-only assumption that genetic variants and environmental exposures are independent in the general population. While this assumption holds naturally in randomized pharmacogenetic studies^47^, additional care is required in observational settings. The current implementation supports such extensions by allowing binary environmental exposures to be analyzed analogously to sex, provided exposure-specific prevalence is available.

In conclusion, we demonstrate that the widely adopted controls-only allelic test by sex is susceptible to systematic type I error inflation because its underlying inferential target is inconsistent with the population quantities relevant to genetic association studies. Rather than representing a flaw in implementation, this bias arises from an implicit assumption that has received little attention despite the widespread use of the method. By reconstructing population-level allele frequencies from both cases and controls, SPADE provides a proper alternative that improves calibration while preserving biologically informative variants. Our work illustrates how re-examining long-standing assumptions underlying established analytical procedures can lead not only to improved statistical methodology but also to new opportunities for biological discovery.

## Data Availability

All data produced in the present work are contained in the manuscript

https://research.mss.ng/

https://base.sfari.org

## Acknowledgements

We thank the families participating in MSSNG, SSC, and SPARK, and acknowledge Autism Speaks and The Centre for Applied Genomics for supporting these resources. Summary statistics derived from these cohorts were used in this study. We thank Michael Escobar and Nancy Reid for helpful discussions. D.Z.C. and C.M. were supported by the CAMH Discovery Fund and the Ontario Brain Institute Centre for Analytics Neuroinformatics Initiative.

M.M.A. was supported by the SickKids Restracomp Fellowship.

## Notes

### Competing Interest Statement

The authors have declared no competing interest.

### Author Declarations

Only aggregated/summary data were used in this study, including population-level genotype counts and summary statistics from previously published sex-specific GWAS. No individual-level data were used, and therefore no de-identification of individual-level data was required.

