## Supplementary file for "Controls-Only Quality Control Metric from Early GWAS Can Attenuate Gene-Sex Interaction Signals in Contemporary Large-Scale Studies"

**Supplementary Note 1. Full derivation for Bias of Allelic Differences by Sex in Controls.**

We start from Bayes' theorem to relate genotype frequencies between cases and the general population:

$$P(\text{ genotype } | \text{ affection status }) = \frac{P(\text{genotype, affection status})}{P(\text{affection status})}$$
$$= P(\text{ affection status } | \text{ genotype }) \cdot \frac{P(\text{genotype})}{P(\text{affection status})}.$$

Here we parameterize the penetrance using a baseline risk and relative risks. Assume allele A is the alternative allele without loss of generality:

$$\begin{aligned} P(\text{ case } | \text{ AA }) &= \kappa_0 \\ P(\text{ case } | \text{ AB }) &= \kappa_0 \cdot \eta_1 \\ P(\text{ case } | \text{ BB }) &= \kappa_0 \cdot \eta_2 \end{aligned}$$

where $\eta_1, \eta_2$ denote genotype-specific relative risks with respect to AA.

$$P(\text{ control } | \text{ genotype }) = 1 - P(\text{ case } | \text{ genotype })$$

The marginal disease prevalence is obtained by averaging over genotype frequencies:

$$\begin{aligned} P(\text{case}) &= P(\text{ case } | \text{ AA })P(\text{AA}) + P(\text{ case } | \text{ AB })P(\text{AB}) + P(\text{ case } | \text{ BB })P(\text{BB}) \\ &= \kappa_0 P(\text{AA}) + \kappa_0\eta_1 P(\text{AB}) + \kappa_0\eta_2 P(\text{BB}) \end{aligned}$$

Solving for $\kappa_0$ gives:

$$\kappa_0 = \frac{P(\text{case})}{P(\text{AA}) + \eta_1 P(\text{AB}) + \eta_2 P(\text{BB})}. \qquad \textbf{S1.1}$$

In males under the non-pseudoautosomal region (NPR), genotypes reduce to single alleles (hemizygous case):

$$\begin{aligned} P(\text{ case } | \text{ A }) &= \kappa_{0,m} \\ P(\text{ case } | \text{ B }) &= \kappa_{0,m} \cdot \eta_{1,m} \end{aligned}$$

Thus, the prevalence simplifies to:

$$P(\text{case}) = \kappa_{0,m} P(\text{A}) + \kappa_{0,m}\eta_{1,m} P(\text{B}),$$

and

$$\kappa_{0,m} = \frac{P(\text{case})}{P(\text{A}) + \eta_{1,m}P(\text{B})}. \quad \textbf{S1.2}$$

We derive the alternative allele frequency among sex s cases:

$$p_{s,a} = P_s(\text{AA|case}) + \frac{1}{2}P_s(\text{AB|case}),$$

where the heterozygote contributes half an allele.
Using Bayes' theorem:

$$p_{s,a} \quad = \kappa_{0,s} \cdot \frac{P_s(\text{AA})}{\pi_s} + \kappa_{0,s} \cdot \eta_{1,s} \cdot \frac{P_s(\text{AB})}{2\pi_s},$$

Substituting S1.1 into the equation and rewriting genotype frequencies in terms of allele frequency $p_s$ and inbreeding coefficient $\delta_s$,

$$P(\text{AA}) = p_s^2 - \delta_s, P(\text{AB}) = 2(p_s(1-p_s) + \delta_s), P(\text{BB}) = (1-p_s)^2 - \delta_s,$$

we obtain:

$$p_{s,a} = \frac{(1-\eta_{1,s})(p_s^2 - \delta_s) + \eta_{1,s}p_s}{(1 - 2\eta_{1,s} + \eta_{2,s})(p_s^2 - \delta_s) + 2(\eta_{1,s} - \eta_{2,s})p_s + \eta_{2,s}}.$$

For males under NPR, the alternative allele frequency among cases is:

$$p_{m,a} = \kappa_{0,m} \cdot \frac{p_m}{\pi_m},$$

Substituting S1.2 into the equation and we get:

$$p_{m,a} = \frac{p_m}{p_m + \eta_{1,m} - \eta_{1,m}p}.$$

We then derive the allele frequency among controls for a sex $s \in \{f, m\}$. By definition,

$$p_{s,u} = P_s(\text{AA|control}) + \frac{1}{2}P_s(\text{AB|control}),$$

Applying Bayes' theorem,

$$p_{s,u} \quad = (1 - \kappa_{0,s}) \cdot \frac{P_s(\text{AA})}{1 - \pi_s} + (1 - \kappa_{0,s} \cdot \eta_{1,s}) \cdot \frac{P_s(\text{AB})}{2(1 - \pi_s)},$$

where $\pi_s = P_s(\text{case})$.

Rearranging to collect allele contributions,

$$p_{s,u} = \frac{(2P_s(\mathrm{AA}) + P_s(\mathrm{AB})) - \kappa_{0,s}\big(2P_s(\mathrm{AA}) + \eta_{1,s}P_s(\mathrm{AB})\big)}{2(1-\pi_s)}.$$

Using $2P(\mathrm{AA}) + P(\mathrm{AB}) = 2p_s$,

$$p_{s,u} = \frac{p_s}{1-\pi_s} - \frac{P_s(\mathrm{AA}) + \eta_{1,s}P_s(\mathrm{AB})/2}{P_s(\mathrm{AA}) + \eta_{1,s}P_s(\mathrm{AB}) + \eta_{2,s}P_s(\mathrm{BB})} \cdot \frac{\pi_s}{1-\pi_s}.$$

Substituting genotype frequencies,

$$p_{s,u} = \frac{p_s}{1-\pi_s} - \frac{(p_s^2-\delta_s) + \eta_{1,s}(2p_s(1-p_s)+2\delta_s)/2}{(p_s^2-\delta_s) + \eta_{1,s}(2p_s(1-p_s)+2\delta_s) + \eta_{2,s}((1-p_s)^2-\delta_s)} \cdot \frac{\pi_s}{1-\pi_s}.$$

After algebraic simplification,

$$\begin{aligned} p_{s,u} &= \frac{p_s}{1-\pi_s} - \frac{(1-\eta_{1,s})p_s^2 + \eta_{1,s}p_s + (\eta_{1,s}-1)\delta_s}{(1-2\eta_{1,s}+\eta_{2,s})p_s^2 + (2\eta_{1,s}-2\eta_{2,s})p_s + \eta_{2,s} + (-1+2\eta_{1,s}-\eta_{2,s})\delta_s} \cdot \frac{\pi_s}{1-\pi_s} \\ &= \frac{p_s}{1-\pi_s} - \frac{(1-\eta_{1,s})(p_s^2-\delta_s) + \eta_{1,s}p_s}{(1-2\eta_{1,s}+\eta_{2,s})(p_s^2-\delta_s) + 2(\eta_{1,s}-\eta_{2,s})p_s + \eta_{2,s}} \cdot \frac{\pi_s}{1-\pi_s}. \end{aligned}$$

Thus, the difference between two sexes $f$ and $m$ is

$$\begin{aligned} p_{f,u} - p_{m,u} &= \left(p_f - \pi_f \cdot \frac{(1-\eta_{1,f})(p_f^2-\delta_f) + \eta_{1,f}p_f}{(1-2\eta_{1,f}+\eta_{2,f})(p_f^2-\delta_f) + 2(\eta_{1,f}-\eta_{2,f})p_f + \eta_{2,f}}\right) \cdot \frac{1}{1-\pi_f} \\ &\quad -\left(p_m - \pi_m \cdot \frac{(1-\eta_{1,m})(p_m^2-\delta_m) + \eta_{1,m}p_m}{(1-2\eta_{1,m}+\eta_{2,m})(p_m^2-\delta_m) + 2(\eta_{1,m}-\eta_{2,m})p_m + \eta_{2,m}}\right) \cdot \frac{1}{1-\pi_m}. \end{aligned}$$

For the X chromosome NPR, males are hemizygous, so the male control allele frequency simplifies to

$$\begin{aligned} p_{m,u} &= P_m(\mathrm{A}|\mathrm{control}) \\ &= (1-\kappa_{0,m}) \cdot \frac{p_m}{1-\pi_m} \\ &= \left(p_m - \pi_m \frac{p_m}{p_m + \eta_{1,m} - \eta_{1,m}p_m}\right) \cdot \frac{1}{1-\pi_m}. \end{aligned}$$

Substituting this into the difference yields

$$p_{f,u} - p_{m,u} = \left(p_f - \pi_f \cdot \frac{(1-\eta_{1,f})(p_f^2 - \delta_f) + \eta_{1,f} p_f}{(1 - 2\eta_{1,f} + \eta_{2,f})(p_f^2 - \delta_f) + 2(\eta_{1,f} - \eta_{2,f}) p_f + \eta_{2,f}}\right) \cdot \frac{1}{1-\pi_f} - \left(p_m - \pi_m \frac{p_m}{p_m + \eta_{1,m} - \eta_{1,m} p_m}\right) \cdot \frac{1}{1-\pi_m}.$$

Combining the above derivations, the difference in allele frequency between female and male controls can be written in a unified form as

$$p_{f,u} - p_{m,u} = \left(p_f - \pi_f \cdot p_{f,a}\right) \cdot \frac{1}{1-\pi_f} - \left(p_m - \pi_m \cdot p_{m,a}\right) \cdot \frac{1}{1-\pi_m}.$$

Here, $p_s$ denotes the population allele frequency for sex $s$, $\pi_s$ the disease prevalence, and $p_{s,a}$ the allele frequency among cases.

### Supplementary Note 2. Extended simulation scenarios for sample size 8000, and power simulation setup.

We extended the simulation to early GWAS setting to show the validity of existing method in smaller sample sizes, as it is underpowered to detect the bias. We simulated data from 4,000 cases and 4,000 controls, a sample size representative of early GWAS. For effect modeling, we considered four genetic architectures for NPR variants and three for autosomal/PAR variants (**Table S1**) consistent with the primary analysis. Three sex prevalence ratios (male: female = 3%:1%, 5%:5%, 1%:3%) were considered. Each scenario was replicated 10,000 times across a range of allele frequencies (0.01, 0.05, 0.1, 0.2, 0.35, 0.5, 0.7, 0.9). Yielding 96 total scenarios for NPR variants and 64 for autosomal variants. We compared both method across a wide range of significance threshold (0.05, 0.01, 0.005, 0.001, 0.0005).

We then simulated power to compare both methods for sample sizes of 40,000 and 400,000 scenarios. For genetic architecture, we considered either a strong main effect alone or no effect (all $\eta_{g,s} = 0$). Two sex prevalence pairs were examined (male: female = 1%:3%, 14%:16%). Three AFs were considered (0.01, 0.1, 0.35). A significance threshold of 1E-6 was used. Three alternative scenarios were evaluated:

1. Population-level sex differences existed; for each AF above, the corresponding male AFs were (0.005, 0.08, 0.032).
2. Measurement error occurred in cases, where 30% of homozygous major genotype frequency was assigned to the homozygous minor genotype frequency for males in cases.
3. Measurement error occurred in controls, where 5% of homozygous

major genotype frequency was assigned to the homozygous minor genotype frequency for males in controls.

**Supplementary Table 1. Genetic architecture parameters across regions.**

| Region | Genetic Architecture | $\eta_{1,f}$ | $\eta_{2,f}$ | $\eta_{1,m}$ | $\eta_{2,m}$ |
|---|---|---|---|---|---|
| X-chromosome non-pseudo-autosomal region | Male-only effect | 1 | 1 | 2 | * |
| | Female-only effect | 1.5 | 2 | 1 | * |
| | Strong main effect | 1.5 | 2 | 2 | * |
| | Opposite-direction | 1.25 | 1.5 | 0.8 | * |
| Autosomal/ pseudo-autosomal region | Single sex effect | 1 | 1 | 1.5 | 2 |
| | Strong main effect | 1.1 | 1.2 | 1.5 | 2 |
| | Opposite-direction | 0.9 | 0.81 | 1.2 | 1.44 |

Note: Asterisk (*) stands for not applicable, $\eta_{g,s}$ is the genotype specific multiplicative factor over base genotype penetrance. Male-only and Female-only effect structure is merged into single sex effective structure to avoid repetition for autosomal region.

**Supplementary Figure 1. Type I error rates for SPADE and control-based QC approaches under a sample size of 8,000, assuming sex-specific disease prevalence (male = 1%, female = 3%) for autosomal variants.**

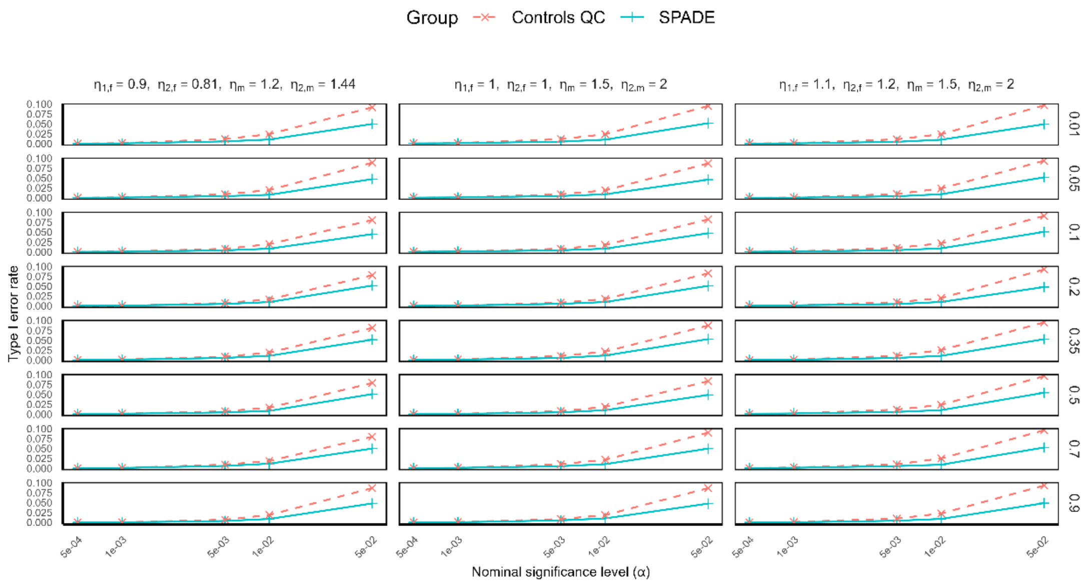


**Supplementary Figure 2. Type I error rates for SPADE and control-based QC approaches under a sample size of 8,000, assuming sex-specific disease prevalence (male = 3%, female = 1%) for autosomal variants.**

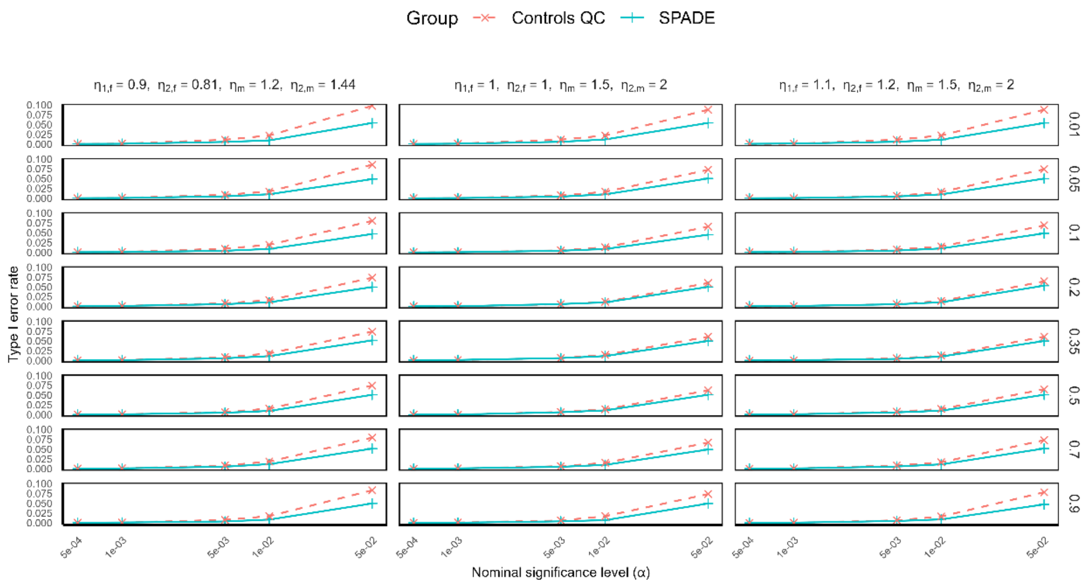

**Supplementary Figure 3. Type I error rates for SPADE and control-based QC approaches under a sample size of 8,000, assuming sex-specific disease prevalence (male = 5%, female = 5%) for autosomal variants.**

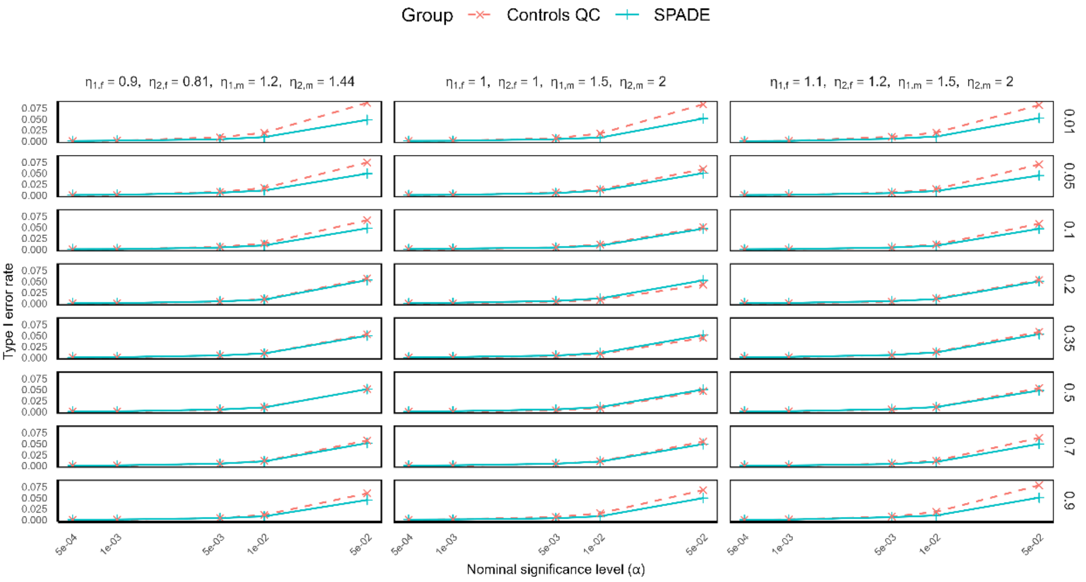


**Supplementary Figure 4. Type I error rates for SPADE and control-based QC approaches under a sample size of 8,000, assuming sex-specific disease prevalence (male = 1%, female = 3%) for NPR variants.**

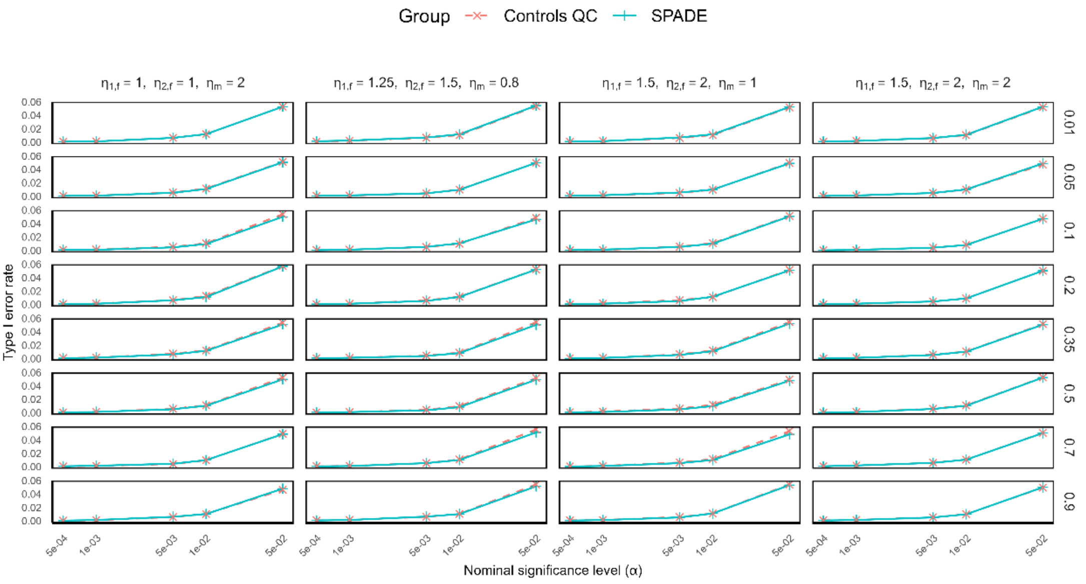

**Supplementary Figure 5. Type I error rates for SPADE and control-based QC approaches under a sample size of 8,000, assuming sex-specific disease prevalence (male = 3%, female = 1%) for NPR variants.**

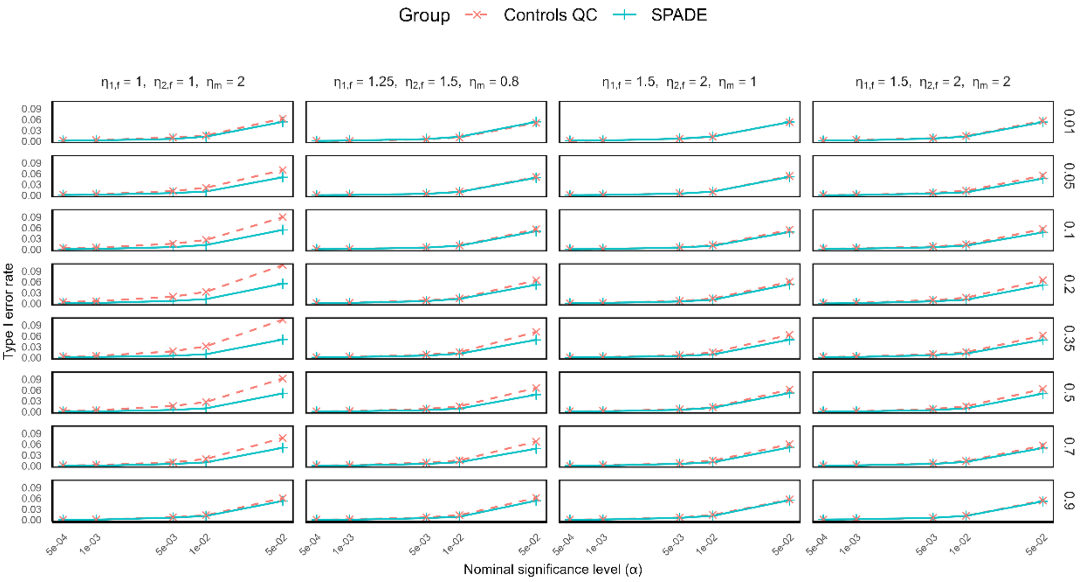


**Supplementary Figure 6. Type I error rates for SPADE and control-based QC approaches under a sample size of 8,000, assuming sex-specific disease prevalence (male = 5%, female = 5%) for NPR variants.**

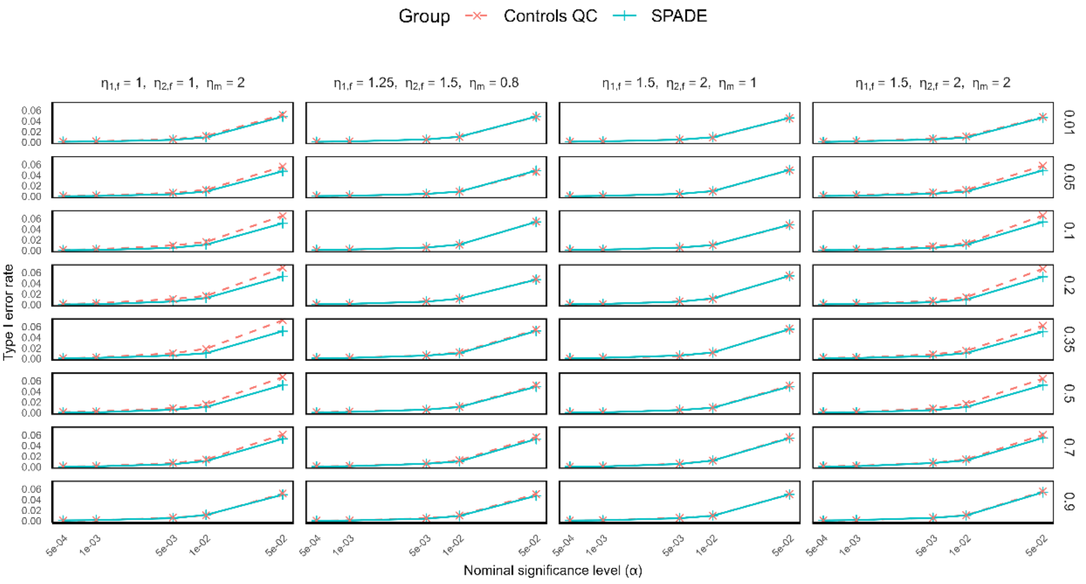

**Supplementary Figure 7. Type I error rate simulation for autosomal/PAR variants.**

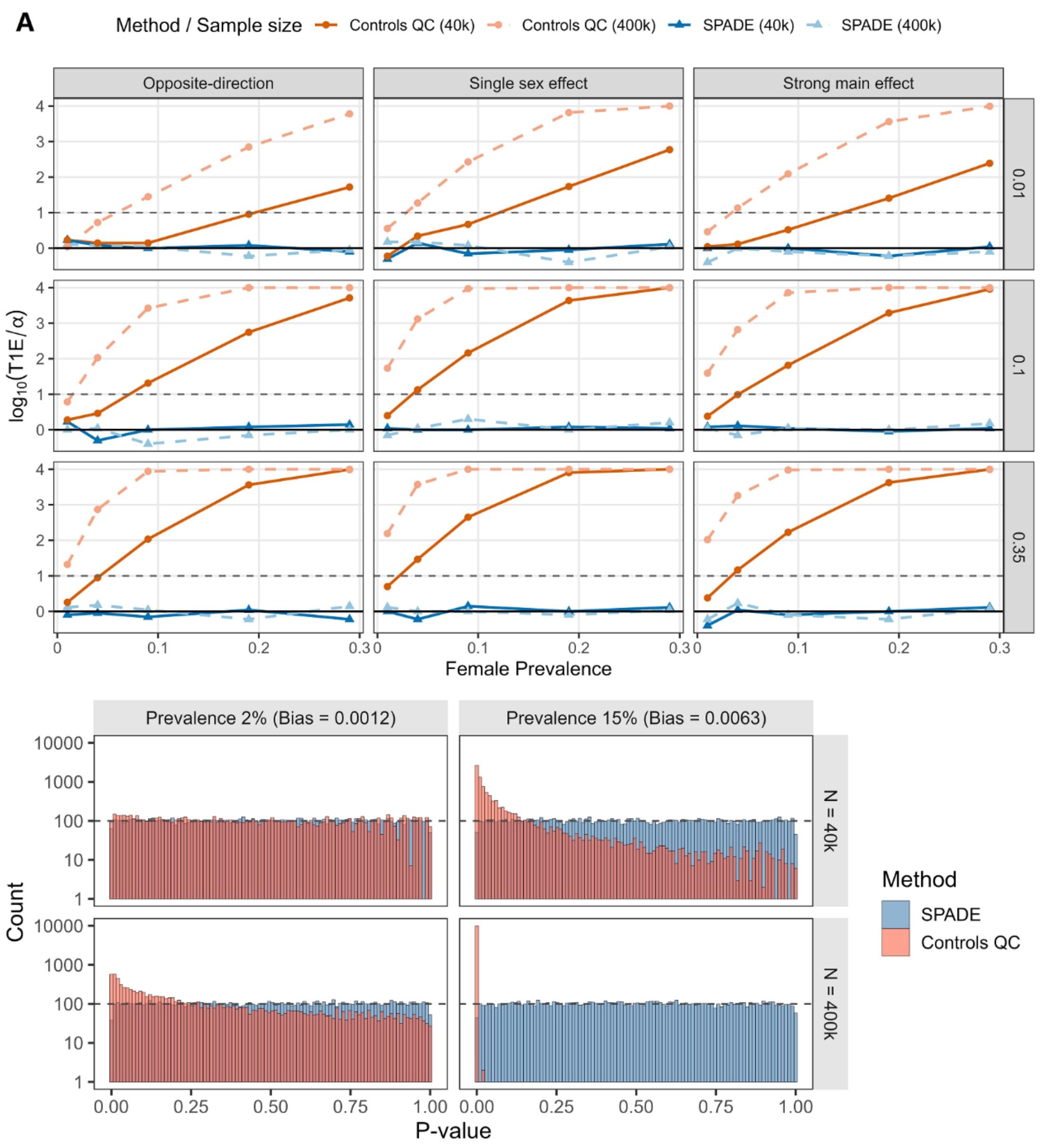


Long legend:

- **(A)** T1E inflation plots across scenarios assessed at $\alpha = 1 \times 10^{-4}$. The facet is stratified by genetic architecture over allele frequencies of (0.01, 0.1, 0.35). The gray dashed line at $y = 1$ represents a 10-fold inflation (i.e. T1E = $1 \times 10^{-3}$), while the zero line represents no inflation.
- **(B)** Histogram of p-value distribution with AF = 0.1 and under a strong main effect genetic architecture. The gray dashed line represents expected density for a uniform(0,1) random variable.

**Supplementary Table 2. Power simulation of SPADE versus controls-only allelic test across scenarios (N = 40,000).**

| $\alpha = 1 \times 10^{-6}$ | | | SNP Associated with the Trait | | | | SNP Unassociated with the Trait | | | |
|---|---|---|---|---|---|---|---|---|---|---|
| | | | Autosome/PAR | | NPR | | Autosome/PAR | | NPR | |
| **Scenario** | **Prevalence** | **AF** | **SPADE** | **Controls-QC** | **SPADE** | **Controls-QC** | **SPADE** | **Controls-QC** | **SPADE** | **Controls-QC** |
| **Population level Sex Differences** | 0.02 | 0.35 | 0.9449 | 0.9751 | 0.6751 | 0.8608 | 0.947 | 0.9295 | 0.6696 | 0.6246 |
| | | 0.1 | 0.9904 | 0.9946 | 0.8576 | 0.9199 | 0.9877 | 0.9828 | 0.846 | 0.8119 |
| | | 0.01 | 0.866 | 0.8592 | 0.5983 | 0.6003 | 0.8518 | 0.8267 | 0.5844 | 0.546 |
| | 0.15 | 0.35 | 0.9944 | 1 | 0.8981 | 0.9997 | 0.9938 | 0.9301 | 0.8766 | 0.622 |
| | | 0.1 | 0.9997 | 1 | 0.9845 | 0.9987 | 0.9993 | 0.9828 | 0.9603 | 0.8119 |
| | | 0.01 | 0.9785 | 0.9289 | 0.8807 | 0.7252 | 0.9711 | 0.8267 | 0.8071 | 0.546 |
| **Error in Cases** | 0.02 | 0.35 | 0 | 0 | 0 | 0 | 0 | 0 | 0 | 0 |
| | | 0.1 | 0.0047 | 0 | 0.002 | 0 | 0.0078 | 0 | 0.0037 | 0 |
| | | 0.01 | 1 | 0 | 0.9972 | 0 | 1 | 0 | 0.9966 | 0 |
| | 0.15 | 0.35 | 0.1083 | 0.0141 | 0.4165 | 0.0469 | 0.509 | 0 | 0.9041 | 0 |
| | | 0.1 | 1 | 0.003 | 1 | 0.0121 | 1 | 0 | 1 | 0 |
| | | 0.01 | 1 | 0 | 1 | 0 | 1 | 0 | 1 | 0 |
| **Error in Controls** | 0.02 | 0.35 | 0.2928 | 0.1615 | 0.7353 | 0.4714 | 0.2895 | 0.3048 | 0.7172 | 0.7319 |
| | | 0.1 | 1 | 1 | 1 | 1 | 1 | 1 | 1 | 1 |
| | | 0.01 | 1 | 1 | 1 | 1 | 1 | 1 | 1 | 1 |
| | 0.15 | 0.35 | 0.333 | 0.0022 | 0.7984 | 0.0131 | 0.2667 | 0.3041 | 0.6929 | 0.7347 |
| | | 0.1 | 1 | 1 | 1 | 1 | 1 | 1 | 1 | 1 |
| | | 0.01 | 1 | 1 | 1 | 1 | 1 | 1 | 1 | 1 |

**Supplementary Table 3. Power simulation of SPADE versus controls-only allelic test across scenarios (N = 400,000).**

| $\alpha = 1 \times 10^{-6}$ | | | SNP Associated with the Trait | | | | SNP Unassociated with the Trait | | | |
|---|---|---|---|---|---|---|---|---|---|---|
| | | | Autosome/PAR | | NPR | | Autosome/PAR | | NPR | |
| **Scenario** | **Prevalence** | **AF** | **SPADE** | **Controls-QC** | **SPADE** | **Controls-QC** | **SPADE** | **Controls-QC** | **SPADE** | **Controls-QC** |
| **Population level Sex Differences** | 0.02 | 0.35 | 1 | 1 | 1 | 1 | 1 | 1 | 1 | 1 |
| | | 0.1 | 1 | 1 | 1 | 1 | 1 | 1 | 1 | 1 |
| | | 0.01 | 1 | 1 | 1 | 1 | 1 | 1 | 1 | 1 |
| | 0.15 | 0.35 | 1 | 1 | 1 | 1 | 1 | 1 | 1 | 1 |
| | | 0.1 | 1 | 1 | 1 | 1 | 1 | 1 | 1 | 1 |
| | | 0.01 | 1 | 1 | 1 | 1 | 1 | 1 | 1 | 1 |
| **Error in Cases** | 0.02 | 0.35 | 0.0016 | 4.00E-04 | 0.007 | 0.0057 | 0.01 | 0 | 0.0502 | 0 |
| | | 0.1 | 0.9895 | 1.00E-04 | 0.9554 | 0.001 | 0.9982 | 0 | 0.9886 | 0 |
| | | 0.01 | 1 | 0 | 1 | 1.00E-04 | 1 | 0 | 1 | 0 |
| | 0.15 | 0.35 | 1 | 0.9995 | 1 | 1 | 1 | 0 | 1 | 0 |
| | | 0.1 | 1 | 0.976 | 1 | 0.9989 | 1 | 0 | 1 | 0 |
| | | 0.01 | 1 | 0.0085 | 1 | 0.0321 | 1 | 0 | 1 | 0 |
| **Error in Controls** | 0.02 | 0.35 | 1 | 1 | 1 | 1 | 1 | 1 | 1 | 1 |
| | | 0.1 | 1 | 1 | 1 | 1 | 1 | 1 | 1 | 1 |
| | | 0.01 | 1 | 1 | 1 | 1 | 1 | 1 | 1 | 1 |
| | 0.15 | 0.35 | 1 | 0.9214 | 1 | 0.9998 | 1 | 1 | 1 | 1 |
| | | 0.1 | 1 | 1 | 1 | 1 | 1 | 1 | 1 | 1 |
| | | 0.01 | 1 | 1 | 1 | 1 | 1 | 1 | 1 | 1 |

**Supplementary Figure 8. Manhattan plot of controls-only QC allele by sex Fisher's exact test.**

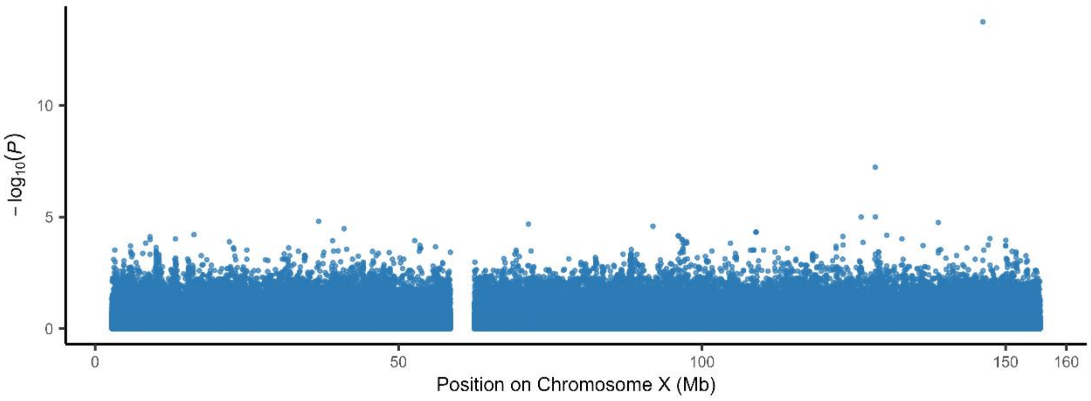


Note: Since Fisher's exact test handles low cell count well, no variant level filter is used, and the above plot covers 418606 variants in total.

**Supplementary Figure 9. Extended QQ-plot across methods.**

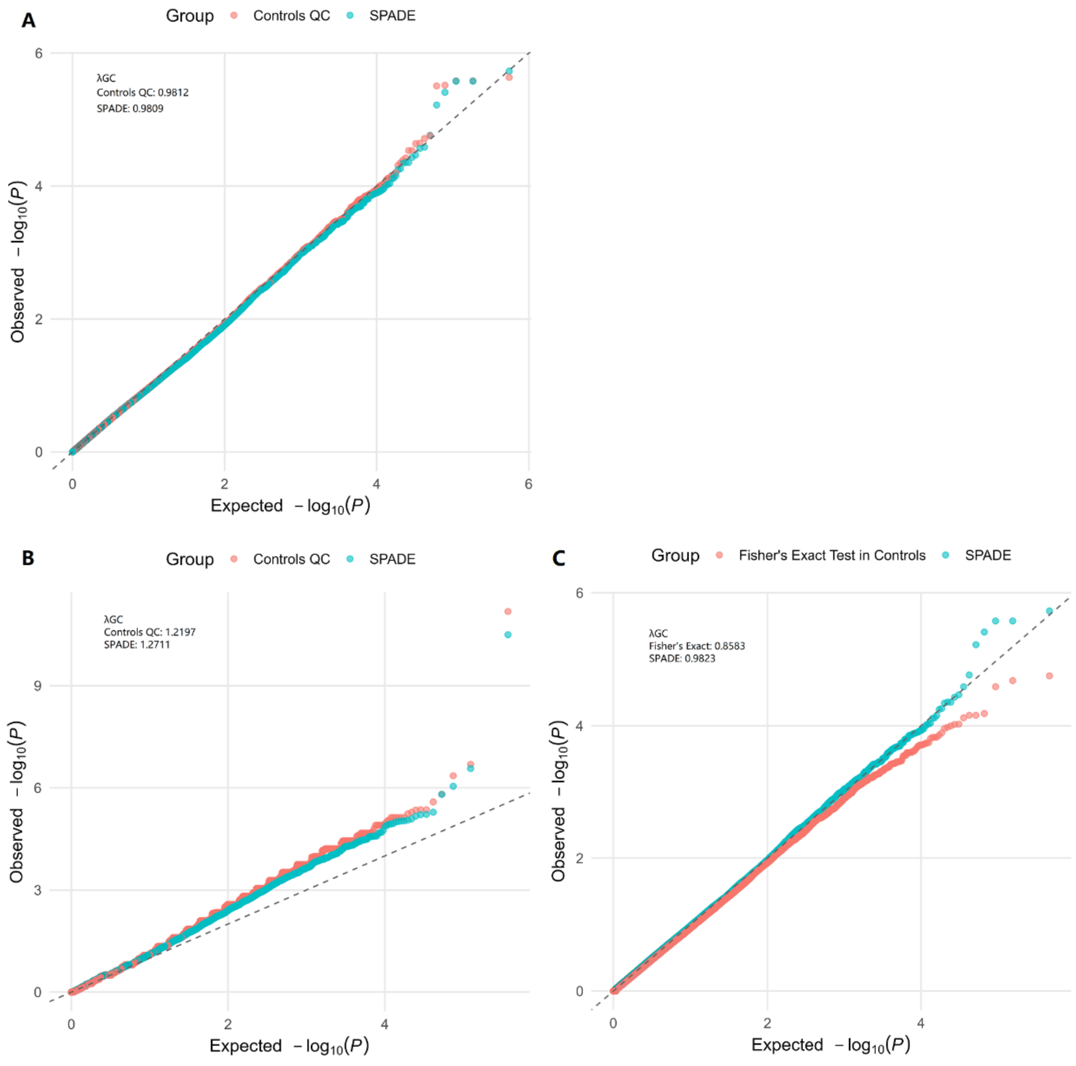

Long legend:

**(A)** QQ-plots by method limited to variants with minor allele count ≥ 5 in controls for both sexes. A total of 279578 variants were plotted. The gray dashed line denotes the identity function (observed = expected).

**(B)** QQ-plots by method limited to variants with minor allele count < 5 in any of affection status by sex stratum. A total of 186384 variants were plotted. The gray dashed line denotes the identity function (observed = expected).

**(C)** QQ-plots by method limited to variants with minor allele count ≥ 5 in any of affection status by sex stratum. A total of 232222 variants were plotted. The gray dashed line denotes the identity function (observed = expected).

**Supplementary Figure 10. Minor allele frequency–stratified QQ-plots comparing Fisher's exact test and SPADE.**

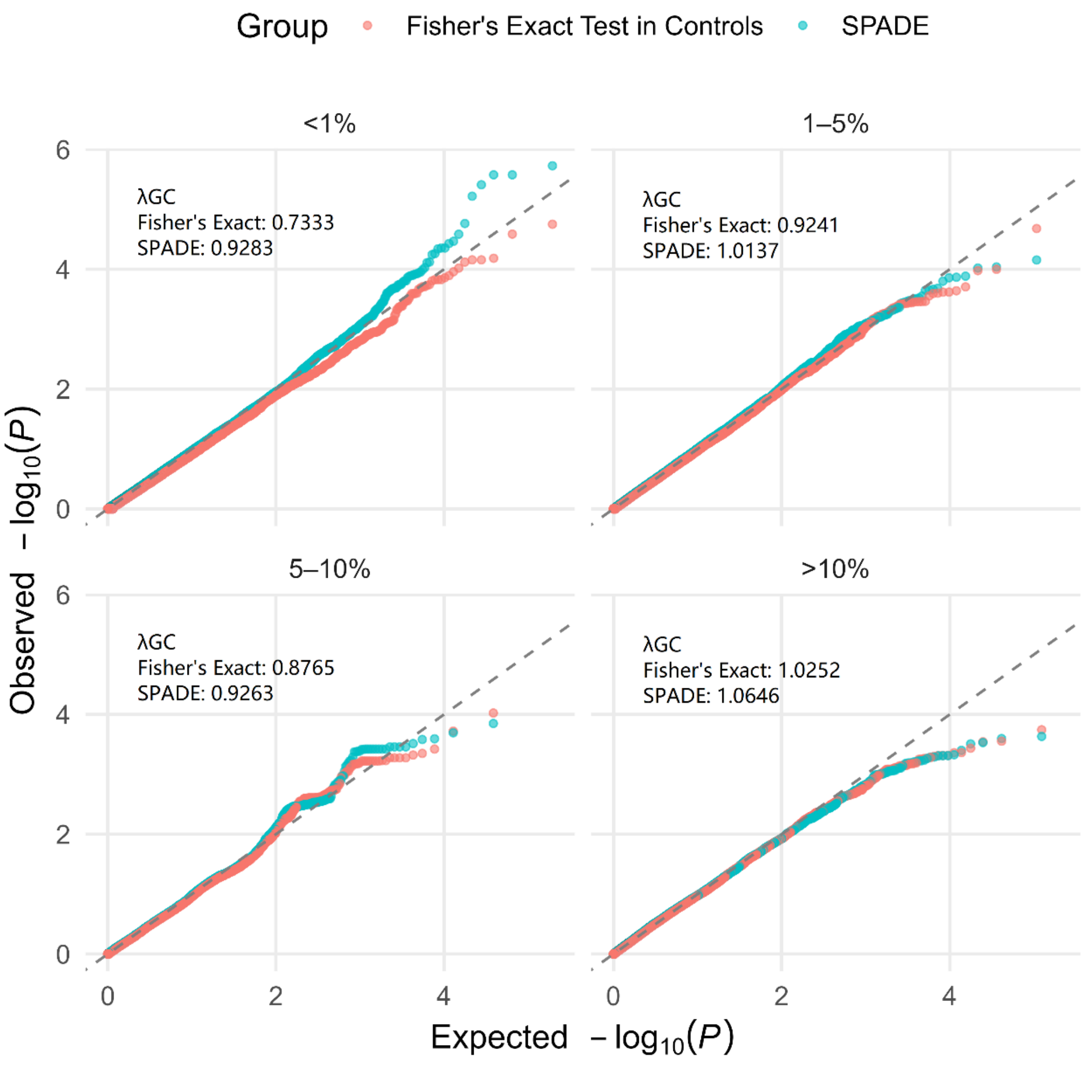


Long legend: Stratified QQ-plots by method limited to variants with minor allele count ≥ 5 in any of affection status by sex stratum. A total of 232222 variants were plotted.

The gray dashed line denotes the identity function (observed = expected).

**Supplementary Figure 11. Minor allele frequency–stratified case–control differences in sex-specific allele frequencies versus the log ratio of female-to-male odds ratios.**

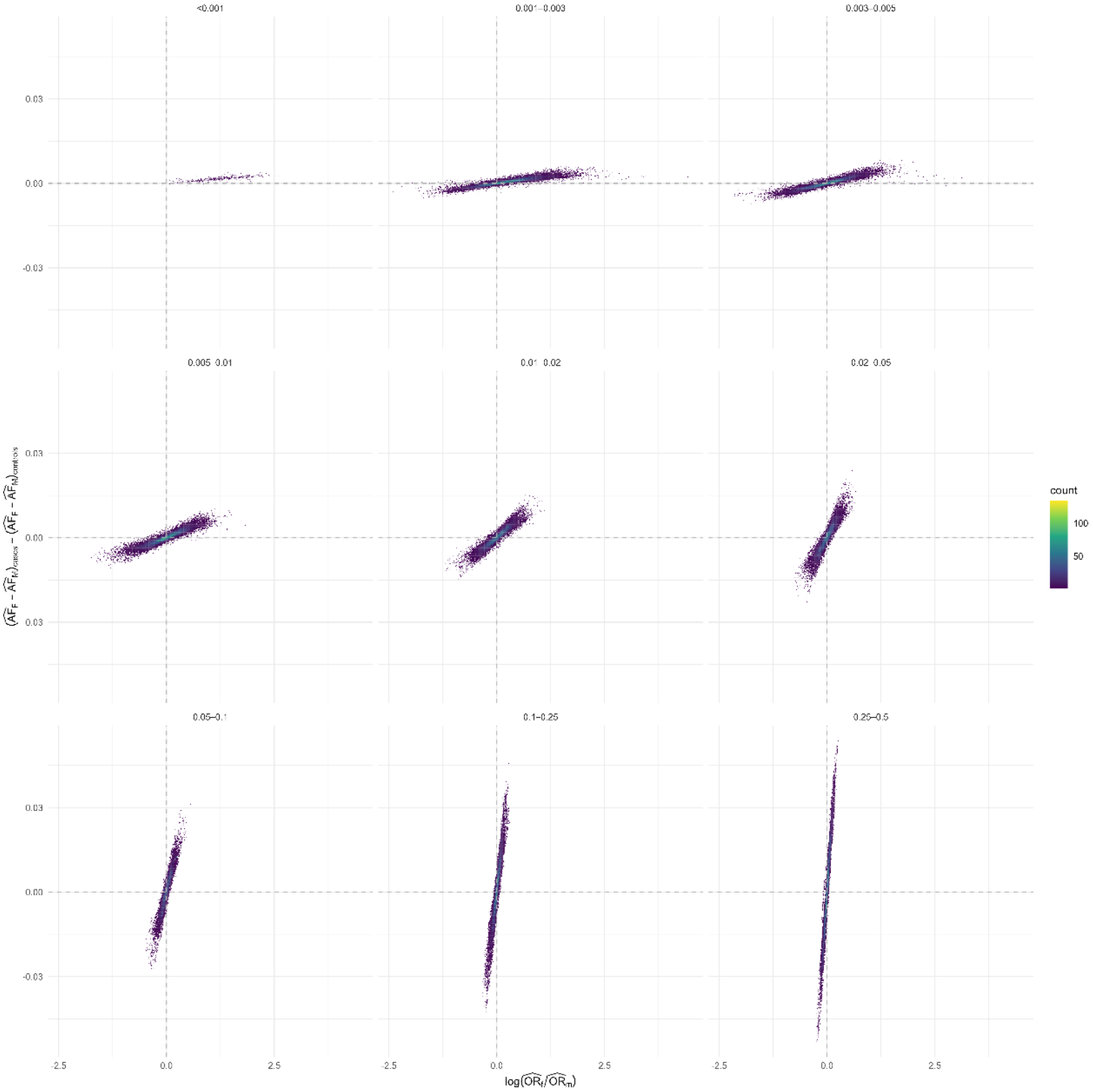
